# An *ex vivo* functional biomarker of treatment response in pediatric low-grade glioma

**DOI:** 10.1101/2025.09.11.25333944

**Authors:** Nichole M Artz, Breanna Mann, Aaron Ebbs, Rami Darawsheh, Rajaneekar Dasari, Adebimpe Adefolaju, Noah Bell, Dimitri Trembath, Dominique Higgins, Scott Elton, Albert Baldwin, Shawn Hingtgen, David E Kram, Andrew B Satterlee

**Affiliations:** Division of Pediatric Hematology-Oncology, Department of Pediatrics, University of North Carolina at Chapel Hill, Chapel Hill, NC, USA; Eshelman School of Pharmacy, Division of Pharmacoengineering and Molecular Pharmaceutics, University of North Carolina at Chapel Hill, Chapel Hill, NC, USA; Eshelman Innovation, University of North Carolina at Chapel Hill, Chapel Hill, NC, USA; Lineberger Comprehensive Cancer Center, University of North Carolina at Chapel Hill, Chapel Hill, NC, USA; Department of Pathology and Laboratory Medicine, University of North Carolina at Chapel Hill, Chapel Hill, NC, USA; Department of Neurosurgery, University of North Carolina at Chapel Hill, Chapel Hill, NC, USA

## Abstract

**Background:** Children with subtotally resected pediatric low-grade glioma (pLGG) often face multiple lines of treatment, which are seldom capable of eliminating the entire tumor. Genomics-based biomarkers are often used to select targeted therapies, but this paradigm only yields overall response rates of ∼50% optimally. Functional precision medicine (FPM), where patient-specific therapeutic efficacy is evaluated by directly treating individuals’ tumor outside their body, can predict individualized drug responses for some cancers, but pLGG is notoriously difficult to maintain outside the body, limiting development of FPM for pLGG.

**Methods:** We describe what is, to our knowledge, the first platform that can maintain, treat, and analyze zero-passage pLGG tumor tissue *ex vivo*, facilitating FPM testing. We engraft pLGG tumors onto a previously validated organotypic brain slice culture (OBSC) platform. After ensuring reproducible engraftment and maintenance of living pLGG tumor tissue on OBSCs, we measured MAPK pathway response to targeted therapies via immunoblotting. We then measured tumor *ex vivo* response to targeted therapies.

**Results:** Each zero-passage pLGG tumor tissue specimen exhibited reproducible growth on the OBSC platform. Western blot demonstrated each BRAF KIAA1549 fusion+ tumor exhibited expected paradoxical MAPK upregulation to dabrafenib treatment. Two of three tumors demonstrated cytotoxicity from trametinib as predicted, whereas one tumor did not. No clinical correlates were measured in this proof-of-concept study, though this mixed response to MEK inhibition may be in line with real-world clinical responses.

**Conclusion:** The OBSC platform supports *ex vivo* maintenance of passage-zero pLGG tumor tissue and enables personalized drug screening to yield a new functional biomarker of pLGG drug response.

## Introduction

Pediatric low-grade gliomas (pLGG) are the most prevalent central nervous system (CNS) tumors in children(1). They are a heterogeneous group of WHO grade 1 and grade 2 tumors arising from glial, neuronal, and mixed glioneuronal cells. While they carry excellent overall survival outcomes(2), they have poor progression-free survival rates, particularly in cases of subtotal resection(3). Patients with incompletely resected pLGG often undergo multiple surgeries and receive several types of therapy over many years, with most therapies failing to destroy the entire tumor. These treatments often lead to debilitating long-term morbidities(4),(5),(6) and severely diminish quality-of-life. To more effectively treat pLGG, limit invasive procedures, and minimize number of treatments given, the field must develop a better method to predict the most effective treatment for each individual’s tumor.

Deficiencies in current biomarkers of response necessitate a change in the way pLGG treatment plans are developed. Currently, many pLGGs are being treated with inhibitors of the mitogen-activated protein kinase (MAPK) pathway, which is most often increased via genetic alterations such as BRAF-V600E mutation and BRAF-KIAA 1549 fusion. Presence of these mutations suggests that kinase inhibitors like dabrafenib (BRAF inhibition) or trametinib (MEK inhibition) may be effective, but only ∼50% of children with BRAF alterations benefit from these targeted therapies. These limitations in genomic tumor biomarkers as predictors of personalized treatment sensitivities are now fueling a shift toward functional precision medicine (FPM). FPM evaluates therapeutic efficacy by directly treating individuals’ living tumor tissue outside their body (after tumor biopsy or resection) to gauge patient-specific activity and response. Some FPM models are beginning to deliver encouraging data to guide matched patient outcomes(7) (8)(9)(10)(11)(12), but pLGG tumor tissues are notoriously difficult to maintain outside the body(13) (14)limiting options to directly screen pLGG tumors for drug sensitivities *ex vivo*.

In this study, we describe what is, to our knowledge, the first platform that can maintain, treat, and analyze passage-zero pLGG tumor tissues *ex vivo*. Here, we engraft passage-zero pLGG tumor tissues onto a previously validated organotypic brain slice culture (OBSC) platform(15). We first ensured reproducible engraftment and maintenance of living pLGG tumor tissue on OBSCs. We then measured maintenance of MAPK pathway responses to dabrafenib and trametinib on OBSCs by assaying the phosphorylation of MEK and ERK via immunoblotting. Finally, we measured sensitivity of each pLGG tumor tissue to dabrafenib and trametinib on OBSCs, correlating *ex vivo* tumor responses to dabrafenib and trametinib to the expected response based on molecular characterization alone.

## Materials and Methods

### Human subjects ethics statement

All brain tumor specimens were collected at University of North Carolina Hospitals. A total of three pediatric patients were included in the present study: two female and one male. Appropriate written informed consent by parent/guardian and assent as applicable was obtained under protocols where ethical approval was given by the Institutional Review Board (IRB) in the Office of Human Research Ethics at the University of North Carolina at Chapel Hill. Two patients consented to non-interventional IRB Protocol #17-2609 (LCCC1805), which was initially approved on 02/15/2018; recruitment is still open as of 09/12/2025. One patient consented to non-interventional IRB Protocol #23-0834 (LCCC2212; ClinicalTrials.gov NCT05978557), which was initially approved on 07/13/2023; recruitment is still open as of 09/12/2025. The laboratory study team had no access to identifying patient information; de-identified or coded diagnostic data describing tumor type and mutational status was provided by a Study Coordinator or Honest Broker.

### Animal ethics statement

All animal work was approved by the Institutional Animal Care and Use Committee at the University of North Carolina at Chapel Hill under protocol 22-171. P8 Sprague-Dawley rat pups were used for all OBSC preparation. Animals were housed with one mother and no more than ten pups. All animals were euthanized prior to weaning.

## METHOD DETAILS

### Chemicals

Dabrafenib (AMBH47A8ED73) and trametinib (AMBH2D6F887E) were purchased from Sigma-Aldrich.

### Lentiviral vectors

mCherry protein fused to firefly luciferase (LV–mCh-FL) were used in this study.

### Generating OBSCs

OBSCs were generated from P8 Sprague-Dawley rat pups using the protocol previously described(15). Briefly, dissected brains were mounted on a vibratome and sliced at a thickness of 300 μm and transferred onto Milicell culture inserts in a 6-well culture plate. 1mL OBSC media was added under each insert and plates were transferred to a 37°C incubator with 5% CO2 and 95% humidity. Readouts were generated a maximum of four days post tumor engraftment.

### Tumor growth on OBSCs

Tumor growth on OBSCs was carried out using the protocol previously described(15). Briefly, pLGG tissue was seeded onto OBSCs 2-hours after slicing, and bioluminescent images obtained 96 hours post-seeding to assess cell viability.

### Drug screening on OBSCs

Drug screening on OBSCs was carried out using the protocol previously described(15). Briefly, tumor cells were seeded onto OBSCs 2-hours after slicing. One day after tumor seeding, drugs were diluted into the media underneath each culture insert. Three days after treatment, bioluminescence of live tumor cells was measured using an AMI optical imager.

### Preparing patient tissue for liquid nitrogen storage

Immediately after brain tumor tissue was surgically resected, tissue was placed in sterile 4°C Neurobasal-A medium and taken to UNC Tissue Procurement Facility (TPF). The resected tumor tissue was minced into approximately 0.5mm pieces by scalpel and washed with PBS, then placed in a cryogenic vial in CryoStor CS10 and frozen at -80°C for 24 hours prior to being transferred into liquid nitrogen.

Immediately after cavitation ultrasonic aspiration tissue was obtained, the manifold containing tissue was placed into a cooler on ice and transported to the laboratory. In a sterile hood, the inner manifold was submerged in 150mL PBS and agitated to release tissue. The tissue was then transferred into 50mL conical tubes and centrifuged for 5 minutes. Once a tissue pellet had formed, supernatant was removed, and 10mL BD Pharm lyse was added to every 2.5mL of combined pellet, incubating at room temperature for 10 minutes. An equal amount of PBS was added to conical tubes containing tissue and RBC lysis buffer and then centrifuged at 1000rpm for 5 minutes. Supernatant was removed and RBC lysis steps repeated. Tissue was transferred onto a 10cm culture dish and manually minced with scalpel for 5 minutes. CryoStor CS10 was added to tissue and divided between cryogenic vials. Cryogenic vials were then frozen at -80°C for 24 hours prior to being transferred into liquid nitrogen.

### Preparing tissue for engraftment onto OBSCs

Preparing tissue for engraftment onto OBSCs was carried out using the protocol previously described(15). Briefly, the preserved brain tumor tissue was thawed, then filtered through 100mm cell strainer and incubated with Lentivirus and polybrene, washed, and reconstituted in Neurobasal-A. The tissue was engrafted onto OBSCs at 0.5mg tissue in 2mL Neurobasal-A and subsequently incubated at 37°C.

### Dosing tissue with drug

Dosing was completed following the standard protocol previously described(15). Briefly, the day after tissue engraftment onto OBSCs, each well in each 6-well plate was dosed with drug. Dose ranges were based on past *in vitro* results(16,17) and OBSC toxicity, and doses were as follows: trametinib ranged from 0 to 100µM, and dabrafenib ranged from 0 to 10µM.Three days after treatment, the bioluminescence of cells was measured using AMI optical imager.

### Western blot analysis

Tumor-bearing OBSCs were snap frozen at 80°C on days 2 and 4 of tumor engraftment. Samples were thawed and suspended in complete lysis buffer (RIPA buffer), then incubated on ice for 5 minutes. Samples were then sonicated in water bath for 5 cycles of 30 seconds on/30 seconds off, then centrifuged at 13,000rpm for 10 minutes at 4°C. Protein concentration was determined in each sample using Bradford assay BioRad Reagent and read on VersaMax plated reader. Protein concentration was then adjusted to 1-2µg/µL in 1x Laemmli Sample Buffer. Samples were heated and run on gel at 120V for 60-90 minutes, then transferred to Nitrocellulose membrane using BioRad turbo blot transfer system. Ponceau solution was used to visualize protein, blots were blocked and agitated for 1 hour at room temperature, then incubated overnight in primary antibody (1:1000) solution at 4°C. Blots were washed a total of 5 times, incubated with secondary antibodies, washed again, and finally, BioRad Clarity ECL solution was used to visualize protein bands on Bio Rad Chemidoc MP imaging system.

### Statistical Analysis

Mean values between two groups were compared using Welch’s two-tailed t tests. The p values are represented as follows: ****p < 0.0001, ***p < 0.001, **p < 0.01, *p ≤ 0.05, and not statistically significant when p > 0.05. The Robust Regression and Outlier Removal (ROUT) method was used to identify outliers from the day 4 microtumor replicates. A Q-value of 1% was used to determine significance. All statistical analyses were performed using GraphPad Prism (version 10.4.1).

## RESULTS

### Tumor Characterization

In this study, tumor tissue was obtained from three pLGG tumors resected between March 2023 and January 2024. The cohort included two female patients and one male patient. Two of the three tumors were gross-totally resected during initial surgery, and none of the patients received tumor-directed therapy following resection. Clinical immunohistochemical analysis confirmed the diagnosis of pLGG, specifically pilocytic astrocytoma, WHO grade 1, for all tumors. BRAF-KIAA 1549 fusion was detected in each tumor for clinical purposes via next generation sequencing customized FusionPlex Solid Tumor kid for Illumina and FusionPlex Supplement for Illumina. None of the tumors harbored the oncogenic BRAF^V600E^ mutation. As of March 2025, at initial manuscript submission, there have been no documented recurrences/progressions. Tumor and patient demographics are listed in Table 1.

**Table 1.** Demographics of pediatric low-grade glioma (pLGG) patients.

| Demographics | pLGG-1 | pLGG-2 | pLGG-3 |
| --- | --- | --- | --- |
| Sex | Male | Female | Female |
| Tumor location | Posterior fossa | Posterior fossa | Posterior fossa |
| Initial resection | Subtotal resection | Gross total resection | Gross total resection |
| Histological diagnosis | Pilocytic astrocytoma<br>CNS WHO grade 1 | Pilocytic astrocytoma<br>CNS WHO grade 1 | Pilocytic astrocytoma<br>CNS WHO grade 1 |
| Genomic alteration | BRAF:KIAA 1549<sup>+</sup> | BRAF:KIAA 1549<sup>+</sup> | BRAF:KIAA 1549<sup>+</sup> |
| Treatment | Surgical resection plus<br>observation | Surgical resection plus<br>observation | Surgical resection plus<br>observation |
| Recurrence | None to date | None to date | None to date |

### Tissue Seeding and Survival on OBSCs

We first tested the reproducible engraftment and survival of all three pLGG tumor tissues on OBSCs. 0.5 mg of mCh-FLuc-transfected pLGG tumor tissue was then seeded atop each hemisphere of freshly prepared, coronally sectioned organotypic brain slice cultures derived from P8 Sprague-Dawley rat pups (Fig 1A). Live tumor cell bioluminescence imaging was performed four days post-seeding and confirmed reproducible pLGG tumor cell survival on OBSCs (Fig 1B). pLGG microtumors were reproducibly established from every seeded replicate, with the only four statistical outliers showing *increased* viability compared to the average survival of each microtumor, possibly due to overseeding. In contrast, pLGG tissue seeded in standard *in vitro* culture showed 43-fold less survival four days after engraftment (p <0.0001 compared to pLGG microtumors on OBSCs, Fig 1C). To our knowledge, this is the first evidence that passage-zero pLGG tumor tissue can be maintained *ex vivo*.

**Figure 1.**
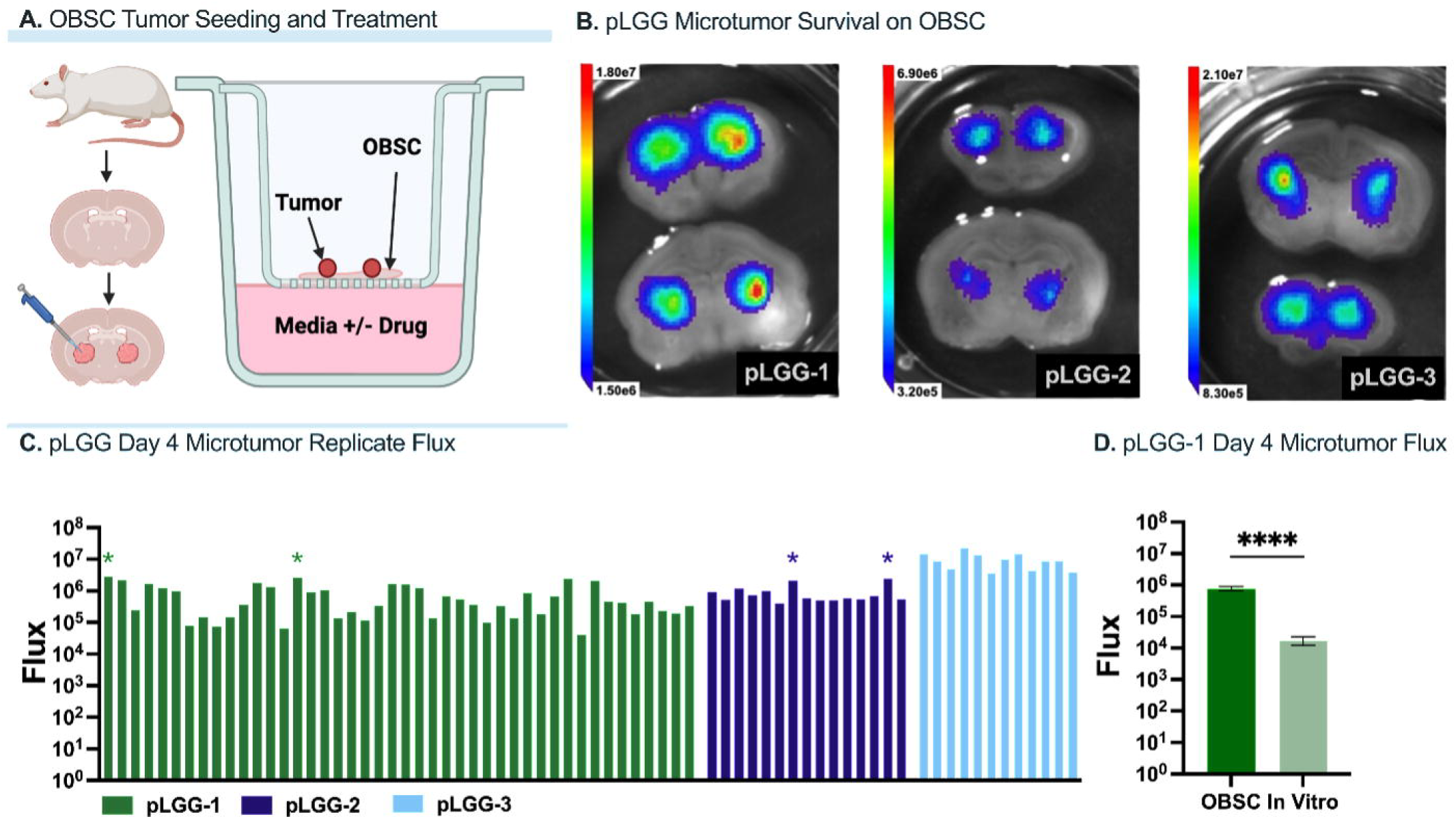
(a) Workflow for organotypic brain slice culture (OBSC) slicing, tumor tissue seeding, and drug treatment. (b) Live tumor cell bioluminescence four days post tumor tissue seeding on OBSCs. (c) pLGG microtumor replicate viability measured via bioluminescent flux, with four statistical outliers showing increased viability compared to the average survival of each microtumor (*statistical outlier measured using ROUT method, Prism). (d) pLGG-1 viability on OBSC compared to in vitro, showing a 43-fold increase in survival on OBSC (****p<0.0001 compared to pLGG-1 microtumors in vitro using Welch’s t test, Prism).

### Western Blot Analysis of Treated pLGG on OBSC

Western blot was performed to assess protein expression of treated pLGG-1 tissue on OBSCs. The BRAF KIAA 1549 fusion event is known to induce a differential effect of MEK inhibition (via treatment with trametinib) and BRAF inhibition (via treatment with dabrafenib) While MEK inhibition leads to deactivation of the MAPK pathway and decreased cell proliferation and survival, BRAF inhibition leads to paradoxical activation of the MAPK pathway due to the binding of the BRAF-monomer inhibitor to one of the dimerized RAF promoters, in turn prohibiting binding to the second RAF promoter through allosteric effects and by decreasing inhibitory phosphorylation. In cells harboring the BRAF KIAA 1549 fusion event, this combination of events leads to increased cell proliferation and survival (Fig 2A)(18,19),(20,21). Here, we treated OBSC-engrafted pLGG-1 with dabrafenib or with trametinib for 24 or 72 hours, then measured phosphorylation levels of MAPK pathway nodes via western blot.

**Figure 2.**
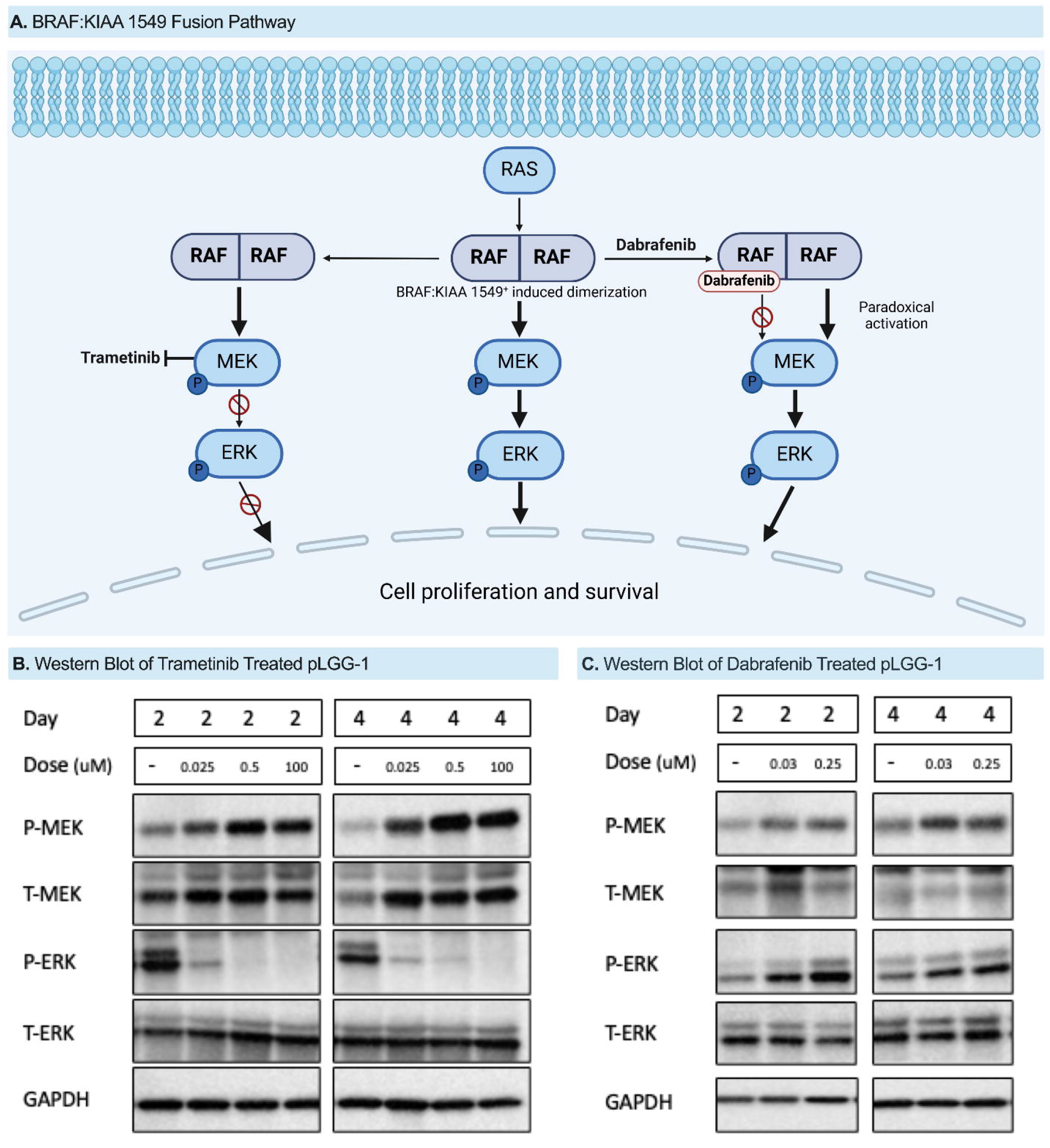
(a) RAS-MAPK pathway demonstrating the effects of the BRAF:KIAA 1549 fusion and targeted therapy with dabrafenib and trametinib. (b) Western blot of BRAF:KIAA 1549+ pLGG-1 cells seeded on organotypic brain slice cultures and treated with trametinib. (c) Western blot of BRAF:KIAA 1549+ pLGG-1 cells seeded on organotypic brain slice cultures and treated with dabrafenib.

As shown in Figure 2B, our results demonstrated a decrease in pERK when the tumor was treated with trametinib and an increase in pERK when the tumor was treated with dabrafenib, consistent with our expectations based on the tumor’s BRAF mutational status. Our results also demonstrated increased pMEK levels after both trametinib and dabrafenib treatment, consistent with our expectations, as both drugs target the MAPK pathway at or upstream of MEK. These data suggest that BRAF-KIAA 1549 fusion^+^ pLGG maintains its molecular phenotype when engrafted on OBSCs, and that this molecular behavior can be specifically measured within the OBSC platform.

### Tumor survival when treated with targeted therapeutics

Finally, we evaluated the responses of pLGG tumor tissues to treatment with dabrafenib and trametinib on OBSCs (Fig 3). Based on all tumors’ BRAF:KIAA1549^+^ status, we expected dabrafenib to be ineffective in all tumors and for trametinib to induce tumor suppression in all tumors – however, historical clinical response data suggests that mutational status does not always predict antitumor efficacy(22). As predicted by the molecular signaling measured in Fig 2, dabrafenib was not effective against pLGG-1, but trametinib induced 68% tumor kill 3 days after treatment (Fig 3A). Similarly, dabrafenib induced tumor growth in pLGG-2 while trametinib induced significant tumor kill (Fig 3B). Interestingly, while dabrafenib was again ineffective against pLGG-3, trametinib was also ineffective, suggesting that other treatment resistance mechanisms may be present in this tumor which could make treatment in the clinical setting less effective (Fig 3C).

**Figure 3.**
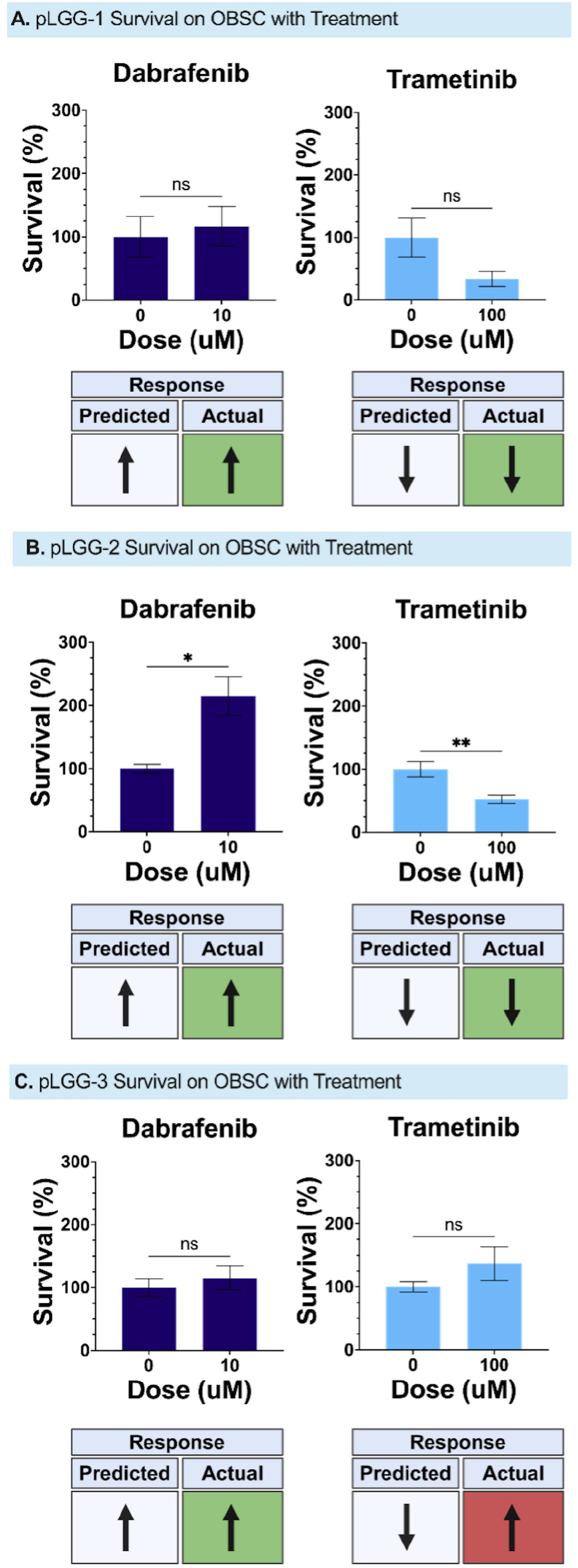
Tumor response to treatment with dabrafenib and trametinib. PLGG-1 (a) and pLGG 2 (b) demonstrated tumor cell growth in response to dabrafenib treatment and tumor cell death in response to trametinib treatment. PLGG-3 (c) demonstrated tumor cell growth in response to dabrafenib and trametinib treatment (*p <0.05, **p<0.01 using Welch’s t test, Prism).

Fortunately for all three of these patients, no treatments were prescribed after surgery. This hindered our ability to correlate our laboratory results with post-treatment patient outcomes, but because progression free survival is as low as 45% with subtotal-resection(23), necessitating effective adjuvant treatment, we believe the data presented here represent a significant step toward validating a functional precision medicine platform for this rare tumor type with few representative *ex vivo* models in existence.

## DISCUSSION

Static, molecular biomarkers such as BRAF-V600E and BRAF-KIAA 1549 fusion alterations are driving treatment guidance in pLGG, but these may not be the only drivers of tumor growth and proliferation within a single patient’s tumor. Tovorafenib and combination dabrafenib and trametinib have both received FDA approval in the past two years(24),(22),(25),(26),(27),(28),(29),(30),(31), and as ongoing clinical trials continue to shed more light on efficacy and safety, especially compared to conventional chemotherapies(32),(33),(34), patients with seemingly identical tumors often experience opposite responses to these targeted agents. To supplement these static biomarkers and better predict a patient’s response to treatment, we describe here a new *functional* biomarker of pLGG response: direct *ex vivo* killing of the tumor tissue itself.

We first demonstrate consistent individualized pLGG tumor engraftment on the OBSC platform. Each of these three patient-derived models demonstrated consistent engraftment and survival. With pLGG-1, the quantity of tissue received enabled us to conduct a greater number of experiments. In this patient-derived pLGG OBSC model, we confirmed the *functional* persistence of the BRAF fusion event by demonstrating downstream MAPK inhibition by a MEK inhibitor and paradoxical downstream upregulation by a first-generation BRAF inhibtor. Finally, and importantly, only 2/3 pLGG tumors in our model responded to trametinib, as measured by cell viability, which more aligns with the clinical experience of incomplete response to MEK across all BRAF KIAA1549^+^ pLGG. However, the greatest strength of the OBSC platform is not in its ability to measure treatment-dependent changes in molecular signaling, but in its ability to quantify whether the tumor ***actually dies***. Tumors like pLGG-3, which do not respond to treatment according to their purported driver mutational status, underscore the importance of functional precision medicine platforms like this OBSC assay.

This method of engrafting pLGG on OBSCs can also have significant implications in preclinical drug development. A major obstacle in translating novel drugs and drug combinations for pLGG remains the fidelity of the limited pLGG preclinical models in existence. Patient-derived pLGG cells are extremely difficult to serially passage in culture or in animals, which has led to the development of innovative but genetically inauthentic patient-derived *TP53* null pLGG models(35),(36),(37), senescence-blocking models(38), and cell lines with uncommon co-occurring mutations such as CDKN2A(39). While potentially useful for early-stage drug discovery(40), these models cannot capture pLGG patient-specific variability.

Furthermore, current preclinical models may overestimate the potency of some therapeutics compared to their clinical response rate. In contrast to near ubiquitous preclinical efficacy data of the second-generation RAF inhibitor, tovorafenib, against BRAF fusion-mutated cell lines and xenograft models(41), approximately 50% of children with relapsed pLGG receiving the drug do not experience an objective response by RAPNO criteria(22). Together, this underscores a need for this OBSC-based *ex vivo* model of pLGG to guide drug development in the preclinical space as well as in the clinic.(2),(13,42),(35),(43),(44),(45)

Our study has limitations. None of the three patients have received any post-surgery chemotherapy, including trametinib or dabrafenib. The opportunity to correlate our *ex vivo* responses to individualized *in vivo* responses would further validate this novel pLGG model, and we continue to consent patients to our actively accruing clinical feasibility study (NCT05978557) to achieve this goal(46). Additionally, while the presence of the BRAF fusion in the tumor specimen was confirmed prior to OBSC engraftment, we did not repeat this test following engraftment. We begin treatment just one day after tumor seeding and complete the assay just three days later, mitigating any chance for genetic drift in culture(15). Lastly, this model lacks an intact blood-brain barrier, the presence of an adaptive immune component, and a blood supply, which may limit the fidelity of the OBSC model. However, the OBSC itself contains innate stromal and immune cells which react to the tumor.(15) We have observed that not only do tumors respond to anatomical features of the OBSCs on which they are engrafted, but the living cells within OBSCs also respond to the presence of tumor.(15) This reciprocal interaction within a living innate neuro-immune microenvironment leads to and may even facilitate increased tumor survival and helps to maintain the genomic profile of the parent tumor.

Together, our findings highlight a novel functional biomarker of pLGG response to treatment, uniquely capturing living, zero-passage, patient-specific pLGG tumor, enabling drug sensitivity testing for therapeutics such as dabrafenib and trametinib in just four days, and yielding reproducible and patient-specific drug sensitivity results to one day inform individualized patient care.

## Acknowledgements

Figures 1 and 3 created in part in BioRender. Artz, N. (2025) https://BioRender.com/m68y745, https://BioRender.com/a63d218

## Authorship

Study design: N.M.A., B.M., D.E.K., and A.B.S. Study implementation and data collection: N.M.A., B.M, R.Dasari., A.E., R.Darwesheh, A.A. Data analysis: N.M.A., B.M., and N.B. Manuscript draft: N.M.A., B.M., D.E.K., and A.B.S. Resources: D.T., D.H., and S.E. Funding acquisition: A.B., D.E.K., A.B.S, and S.H. Project Administration: A.B., D.E.K., A.B.S, and S.H. All authors have contributed to the manuscript and approved the submitted version.

## Data Availability

All relevant data are within the manuscript.

## References

1. Ostrom QT, Price M, Ryan K, Edelson J, Neff C, Cioffi G, et al. CBTRUS Statistical Report: Pediatric Brain Tumor Foundation Childhood and Adolescent Primary Brain and Other Central Nervous System Tumors Diagnosed in the United States in 2014-2018. Neuro Oncol. 2022 Sep 6;24(Suppl 3):iii1–38.

2. Fangusaro J, Jones DT, Packer RJ, Gutmann DH, Milde T, Witt O, et al. Pediatric low-grade glioma: State-of-the-art and ongoing challenges. Neuro Oncol. 2024 Jan 5;26(1):25–37.

3. Kameda-Smith MM, Green K, Hutton DL, Jeelani NUO, Thompson DNP, Hargrave D, et al. The role of reoperation in pediatric cerebellar pilocytic astrocytoma. J Neurosurg Pediatr. 2024 Aug 1;34(2):169–75.

4. Bandopadhayay P, Bergthold G, London WB, Goumnerova LC, Morales La Madrid A, Marcus KJ, et al. Long-term outcome of 4,040 children diagnosed with pediatric low-grade gliomas: an analysis of the Surveillance Epidemiology and End Results (SEER) database. Pediatr Blood Cancer. 2014 Jul;61(7):1173–9.

5. Krishnatry R, Zhukova N, Guerreiro Stucklin AS, Pole JD, Mistry M, Fried I, et al. Clinical and treatment factors determining long-term outcomes for adult survivors of childhood low-grade glioma: A population-based study. Cancer. 2016 Apr 15;122(8):1261–9.

6. Zelt S, Cooney T, Yu S, Daral S, Krebs B, Markan R, et al. Disease burden and healthcare utilization in pediatric low-grade glioma: A United States retrospective study of linked claims and electronic health records. Neurooncol Pract. 2024 Oct;11(5):583–92.

7. Malani D, Kumar A, Brück O, Kontro M, Yadav B, Hellesøy M, et al. Implementing a functional precision medicine tumor board for acute myeloid leukemia. Cancer Discov. 2022 Feb;12(2):388–401.

8. Kornauth C, Pemovska T, Vladimer GI, Bayer G, Bergmann M, Eder S, et al. Functional precision medicine provides clinical benefit in advanced aggressive hematologic cancers and identifies exceptional responders. Cancer Discov. 2022 Feb;12(2):372–87.

9. Shuford S, Wilhelm C, Rayner M, Elrod A, Millard M, Mattingly C, et al. Prospective Validation of an Ex Vivo, Patient-Derived 3D Spheroid Model for Response Predictions in Newly Diagnosed Ovarian Cancer. Sci Rep. 2019 Aug 1;9(1):11153.

10. Peterziel H, Jamaladdin N, ElHarouni D, Gerloff XF, Herter S, Fiesel P, et al. Drug sensitivity profiling of 3D tumor tissue cultures in the pediatric precision oncology program INFORM. NPJ Precis Oncol. 2022 Dec 27;6(1):94.

11. Mayoh C, Mao J, Xie J, Tax G, Chow S-O, Cadiz R, et al. High-Throughput Drug Screening of Primary Tumor Cells Identifies Therapeutic Strategies for Treating Children with High-Risk Cancer. Cancer Res. 2023 Aug 15;83(16):2716–32.

12. Acanda De La Rocha AM, Berlow NE, Fader M, Coats ER, Saghira C, Espinal PS, et al. Feasibility of functional precision medicine for guiding treatment of relapsed or refractory pediatric cancers. Nat Med. 2024 Apr 11;30(4):990–1000.

13. Milde T, Fangusaro J, Fisher MJ, Hawkins C, Rodriguez FJ, Tabori U, et al. Optimizing preclinical pediatric low-grade glioma models for meaningful clinical translation. Neuro Oncol. 2023 Nov 2;25(11):1920–31.

14. Yvone GM, Breunig JJ. Pediatric low-grade glioma models: advances and ongoing challenges. Front Oncol. 2023;13:1346949.

15. Mann B, Zhang X, Bell N, Adefolaju A, Thang M, Dasari R, et al. A living ex vivo platform for functional, personalized brain cancer diagnosis. Cell Rep Med. 2023 Jun 20;4(6):101042.

16. Witkowski J, Polak S, Rogulski Z, Pawelec D. In vitro/in vivo translation of synergistic combination of MDM2 and MEK inhibitors in melanoma using PBPK/PD modelling: part I. Int J Mol Sci. 2022 Oct 26;23(21).

17. Cordaro FG, De Presbiteris AL, Camerlingo R, Mozzillo N, Pirozzi G, Cavalcanti E, et al. Phenotype characterization of human melanoma cells resistant to dabrafenib. Oncol Rep. 2017 Nov;38(5):2741–51.

18. Holderfield M, Nagel TE, Stuart DD. Mechanism and consequences of RAF kinase activation by small-molecule inhibitors. Br J Cancer. 2014 Aug 12;111(4):640–5.

19. Hanrahan AJ, Chen Z, Rosen N, Solit DB. BRAF - a tumour-agnostic drug target with lineage-specific dependencies. Nat Rev Clin Oncol. 2024 Mar;21(3):224–47.

20. Boussemart L, Girault I, Malka-Mahieu H, Mateus C, Routier E, Rubington M, et al. Secondary Tumors Arising in Patients Undergoing BRAF Inhibitor Therapy Exhibit Increased BRAF-CRAF Heterodimerization. Cancer Res. 2016 Mar 15;76(6):1476–84.

21. Gibney GT, Messina JL, Fedorenko IV, Sondak VK, Smalley KSM. Paradoxical oncogenesis--the long-term effects of BRAF inhibition in melanoma. Nat Rev Clin Oncol. 2013 Jul;10(7):390–9.

22. Kilburn LB, Khuong-Quang D-A, Hansford JR, Landi D, van der Lugt J, Leary SES, et al. The type II RAF inhibitor tovorafenib in relapsed/refractory pediatric low-grade glioma: the phase 2 FIREFLY-1 trial. Nat Med. 2024 Jan;30(1):207–17.

23. Wisoff JH, Sanford RA, Heier LA, Sposto R, Burger PC, Yates AJ, et al. Primary neurosurgery for pediatric low-grade gliomas: a prospective multi-institutional study from the Children’s Oncology Group. Neurosurgery. 2011 Jun;68(6):1548–54; discussion 1554.

24. Bouffet E, Hansford JR, Garrè ML, Hara J, Plant-Fox A, Aerts I, et al. Dabrafenib plus Trametinib in Pediatric Glioma with BRAF V600 Mutations. N Engl J Med. 2023 Sep 21;389(12):1108–20.

25. Ater JL, Zhou T, Holmes E, Mazewski CM, Booth TN, Freyer DR, et al. Randomized study of two chemotherapy regimens for treatment of low-grade glioma in young children: a report from the Children’s Oncology Group. J Clin Oncol. 2012 Jul 20;30(21):2641–7.

26. Bouffet E, Jakacki R, Goldman S, Hargrave D, Hawkins C, Shroff M, et al. Phase II study of weekly vinblastine in recurrent or refractory pediatric low-grade glioma. J Clin Oncol. 2012 Apr 20;30(12):1358–63.

27. Banerjee A, Jakacki RI, Onar-Thomas A, Wu S, Nicolaides T, Young Poussaint T, et al. A phase I trial of the MEK inhibitor selumetinib (AZD6244) in pediatric patients with recurrent or refractory low-grade glioma: a Pediatric Brain Tumor Consortium (PBTC) study. Neuro Oncol. 2017 Aug 1;19(8):1135–44.

28. Perreault S, Larouche V, Tabori U, Hawkin C, Lippé S, Ellezam B, et al. A phase 2 study of trametinib for patients with pediatric glioma or plexiform neurofibroma with refractory tumor and activation of the MAPK/ERK pathway: TRAM-01. BMC Cancer. 2019 Dec 27;19(1):1250.

29. Fangusaro J, Onar-Thomas A, Young Poussaint T, Wu S, Ligon AH, Lindeman N, et al. Selumetinib in paediatric patients with BRAF-aberrant or neurofibromatosis type 1-associated recurrent, refractory, or progressive low-grade glioma: a multicentre, phase 2 trial. Lancet Oncol. 2019 Jul;20(7):1011–22.

30. Bouffet E, Geoerger B, Moertel C, Whitlock JA, Aerts I, Hargrave D, et al. Efficacy and Safety of Trametinib Monotherapy or in Combination With Dabrafenib in Pediatric BRAF V600-Mutant Low-Grade Glioma. J Clin Oncol. 2023 Jan 20;41(3):664–74.

31. Hargrave DR, Bouffet E, Tabori U, Broniscer A, Cohen KJ, Hansford JR, et al. Efficacy and Safety of Dabrafenib in Pediatric Patients with BRAF V600 Mutation-Positive Relapsed or Refractory Low-Grade Glioma: Results from a Phase I/IIa Study. Clin Cancer Res. 2019 Dec 15;25(24):7303–11.

32. CLINICAL TRIAL / NCT03871257 - UChicago Medicine [Internet]. [cited 2025 Jan 21]. Available from: https://www.uchicagomedicine.org/find-a-clinical-trial/clinical-trial/cirb201848

33. Toure A, Ghione P, Phillips S, Klute K, Leonard JP, Martin P. Lymphoma study titles on clinicaltrials.gov lack details necessary for study identification. Clin Lymphoma Myeloma Leuk. 2020 Feb;20(2):e82–6.

34. van Tilburg CM, Kilburn LB, Crotty E, Smith AA, Perreault S, Franson AF, et al. LOGGIC/FIREFLY-2: A phase 3, randomized trial of tovorafenib vs. chemotherapy in pediatric and young adult patients with newly diagnosed low-grade glioma harboring an activating RAF alteration. JCO. 2023 Jun 1;41(16_suppl):TPS10067–TPS10067.

35. Selt F, Hohloch J, Hielscher T, Sahm F, Capper D, Korshunov A, et al. Establishment and application of a novel patient-derived KIAA1549:BRAF-driven pediatric pilocytic astrocytoma model for preclinical drug testing. Oncotarget. 2017 Feb 14;8(7):11460–79.

36. Selt F, Sigaud R, Valinciute G, Sievers P, Zaman J, Alcon C, et al. BH3 mimetics targeting BCL-XL impact the senescent compartment of pilocytic astrocytoma. Neuro Oncol. 2023 Apr 6;25(4):735–47.

37. Bax DA, Little SE, Gaspar N, Perryman L, Marshall L, Viana-Pereira M, et al. Molecular and phenotypic characterisation of paediatric glioma cell lines as models for preclinical drug development. PLoS ONE. 2009 Apr 14;4(4):e5209.

38. Yuan M, White D, Resar L, Bar E, Groves M, Cohen A, et al. Conditional reprogramming culture conditions facilitate growth of lower-grade glioma models. Neuro Oncol. 2021 May 5;23(5):770–82.

39. Bid HK, Kibler A, Phelps DA, Manap S, Xiao L, Lin J, et al. Development, characterization, and reversal of acquired resistance to the MEK1 inhibitor selumetinib (AZD6244) in an in vivo model of childhood astrocytoma. Clin Cancer Res. 2013 Dec 15;19(24):6716–29.

40. Sigaud R, Rösch L, Gatzweiler C, Benzel J, von Soosten L, Peterziel H, et al. The first-in-class ERK inhibitor ulixertinib shows promising activity in mitogen-activated protein kinase (MAPK)-driven pediatric low-grade glioma models. Neuro Oncol. 2023 Mar 14;25(3):566–79.

41. Sun Y, Alberta JA, Pilarz C, Calligaris D, Chadwick EJ, Ramkissoon SH, et al. A brain-penetrant RAF dimer antagonist for the noncanonical BRAF oncoprotein of pediatric low-grade astrocytomas. Neuro Oncol. 2017 Jun 1;19(6):774–85.

42. Pan Y, Xiong M, Chen R, Ma Y, Corman C, Maricos M, et al. Athymic mice reveal a requirement for T-cell-microglia interactions in establishing a microenvironment supportive of Nf1 low-grade glioma growth. Genes Dev. 2018 Apr 1;32(7–8):491–6.

43. Guo X, Pan Y, Xiong M, Sanapala S, Anastasaki C, Cobb O, et al. Midkine activation of CD8+ T cells establishes a neuron-immune-cancer axis responsible for low-grade glioma growth. Nat Commun. 2020 May 1;11(1):2177.

44. de Andrade Costa A, Chatterjee J, Cobb O, Cordell E, Chao A, Schaeffer S, et al. Immune deconvolution and temporal mapping identifies stromal targets and developmental intervals for abrogating murine low-grade optic glioma formation. Neurooncol Adv. 2022;4(1):vdab194.

45. Kolb EA, Gorlick R, Houghton PJ, Morton CL, Neale G, Keir ST, et al. Initial testing (stage 1) of AZD6244 (ARRY-142886) by the Pediatric Preclinical Testing Program. Pediatr Blood Cancer. 2010 Oct;55(4):668–77.

46. Brain Slice Explants to Predict Drug Response in Brain Tumors - National Brain Tumor Society [Internet]. [cited 2025 Jan 21]. Available from: https://trials.braintumor.org/trials/NCT05978557

